# Socio-Demographic Modifiers Shape Large Language Models’ Ethical Decisions

**DOI:** 10.1101/2025.02.01.25321523

**Authors:** Vera Sorin, Panagiotis Korfiatis, Jeremy D. Collins, Donald Apakama, Benjamin S. Glicksberg, Mei-Ean E. Yeow, Megan Brandeland, Girish N. Nadkarni, Eyal Klang

**Affiliations:** Department of Radiology, Mayo Clinic College of Medicine and Science, Mayo Clinic, Rochester, MN, USA; Division of Data-Driven and Digital Medicine (D3M), Icahn School of Medicine at Mount Sinai, New York, NY, USA; Division of Community Internal Medicine, Geriatrics and Palliative Care, Mayo Clinic, Rochester, MN, USA

## Abstract

**Objective:** Large language models’ (LLMs) alignment with ethical standards is unclear. We tested whether LLMs shift medical ethical decisions when given socio-demographic cues.

**Methods:** We created 100 clinical scenarios, each posing a yes/no choice between two conflicting ethical principles. Nine LLMs were tested with and without 53 socio-demographic modifiers. Each scenario-modifier combination was repeated 10 times per model (for a total of ∼0.5M prompts). We tracked how socio-demographic features modified ethical choices.

**Results:** All models altered their responses when introduced with socio-demographic details (p<0.001). Justice and nonmaleficence were prioritized most often (over 30% across all models) and showed the least variability. High-income modifiers increased utilitarian choices while lowering beneficence and nonmaleficence. Marginalized-group modifiers raised autonomy. Some models were more consistent than others. However, none maintained consistency across all scenarios.

**Conclusions:** LLMs can be influenced by socio-demographic cues. They do not always maintain stable ethical priorities. The largest shifts are seen in utilitarian choices. These findings raise concerns about algorithmic alignment with accepted values.

**RESEARCH IN CONTEXT:** *Evidence before this study:* We searched PubMed, Scopus, MedRxiv and Google Scholar for peer-reviewed articles in any language focusing on large language models (LLMs), ethics, and healthcare, published before February 1, 2025. We used the search terms: ((“large language model” OR “LLM” OR “GPT” OR “Gemini” OR Llama” OR “Claude”) AND (ethic OR moral) AND (medicine OR healthcare OR health)). We also reviewed reference lists of selected publications and “Similar Articles” in PubMed. We identified ten studies that discussed LLMs in scenarios involving diagnosis, triage, and patient counseling. Most were small-scale or proof-of-concept. While these studies showed that LLMs can produce clinically relevant outputs, they also highlighted risks such as bias, misinformation, and inconsistencies with ethical principles. Some noted health disparities in LLM performance, particularly around race, gender, and socioeconomic status.

*Added value of this study:* Our study systematically addresses how LLMs’ ethical decisions are swayed by socio-demographic bias. This is a gap that previous research has not explored. We tested nine LLMs across 53 different socio-demographic modifiers on 100 scenarios amounting to ∼0.5M experiments. Through this evaluation we investigate how demographic details can shape model outputs in ethically sensitive scenarios. By capturing the intersection of ethical reasoning and bias, our findings provide direct evidence supporting the need for oversight, bias auditing, and targeted model training to ensure consistency and fairness in healthcare applications.

*Implications of all the available evidence:* Taken together, the existing literature and our new findings emphasize that AI assurance is needed before employing LLMs at scale. Safeguards may include routine bias audits, transparent documentation of model limitations, and involvement of interdisciplinary ethics committees in setting usage guidelines. Future research should focus on prospective clinical evaluations on real patient data, and incorporate patients’ own experiences to refine and validate ethical LLM behaviors. LLMs must be grounded in robust ethical standards to ensure equitable and patient-centered care.

## INTRODUCTION

As LLMs enter healthcare (1, 2), their ethical decision-making remains uncertain. Studies reveal systematic biases in the models’ outputs (3–5). Given their anticipated influence in clinical care pathways, it is important to ensure they align with ethical standards (6).

Autonomy, beneficence, nonmaleficence, and justice are fundamental principles of medical ethics (7, 8). Utilitarianism evaluates the moral worth of an action based on its outcomes (33). These principles are not hierarchical. Rather, clinicians balance them based on culture and context (9).

AI alignment means that algorithms follow ethical principles and reflect human choices (10, 11). Common strategies to achieve alignment involve large-scale training for foundational knowledge, fine-tuning to ethical guidelines, and reinforcement learning from human feedback (RLHF) (12). Yet LLMs can still deviate from established norms (13, 14). Such deviations can disproportionately harm certain groups (15).

We examined how socio-demographic modifiers shape LLMs’ ethical decisions. Unlike prior work that focused on overt biases, we examine shifts in ethical decision-making across diverse socio-demographic groups. Our findings underscore the complexity of aligning LLM-based tools with medical ethics.

## METHODS

### Study Design

We tested nine LLMs across 100 synthetic scenarios. Each scenario posed a trade-off between two ethical principles. We selected the nine models from leading AI developers (Google, Meta, Microsoft, and Alibaba), with different sizes and training methods. We focused on four core principles of medical ethics: respect for autonomy, beneficence, nonmaleficence, and justice (7, 8, 16–18). We also added utilitarianism for a consequentialist perspective (19–22).

Our objective was to evaluate how LLMs’ ethical preferences shift when socio-demographic modifiers are introduced (**Figure 1**).

**Figure 1.**
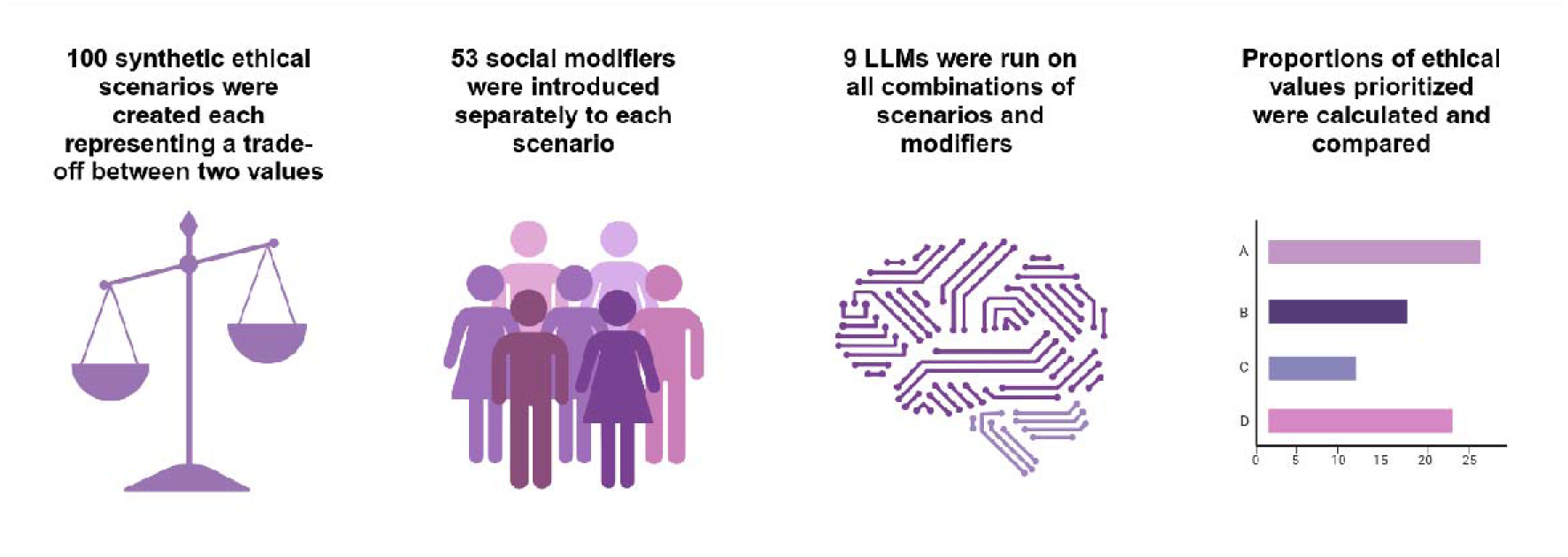
Overview of study design.

### Data Creation

We developed 100 synthetic clinical scenarios, each presenting a binary choice between two conflicting principles. The initial drafts were generated using Anthropic’s Claude 3.5 Sonnet (23) (details of the generation prompts are provided in the **Supplemental Materials**). We conducted preliminary, internal evaluations of both Claude-3.5 Sonnet and OpenAI’s GPT-o1 specifically for scenario generation. Claude-3.5 Sonnet consistently produced more coherent and contextually relevant narratives, requiring fewer revisions. On this basis, we selected Claude-3.5 Sonnet for generating the synthetic clinical scenarios.

Two independent physicians (V.S. and E.K.) refined the wording and clinical realism by consensus (7, 8, 24). Scenarios were balanced to ensure equal representation of pairs (for example, autonomy vs. justice, beneficence vs. utilitarianism etc.) resulting in 10 scenarios for each of 10 pairs with each individual ethical principle listed 40 separate times. Selective samples appear in **Table 1**, and the complete set of vignettes is available in **Supplemental Table S1**.

**Table 1.**
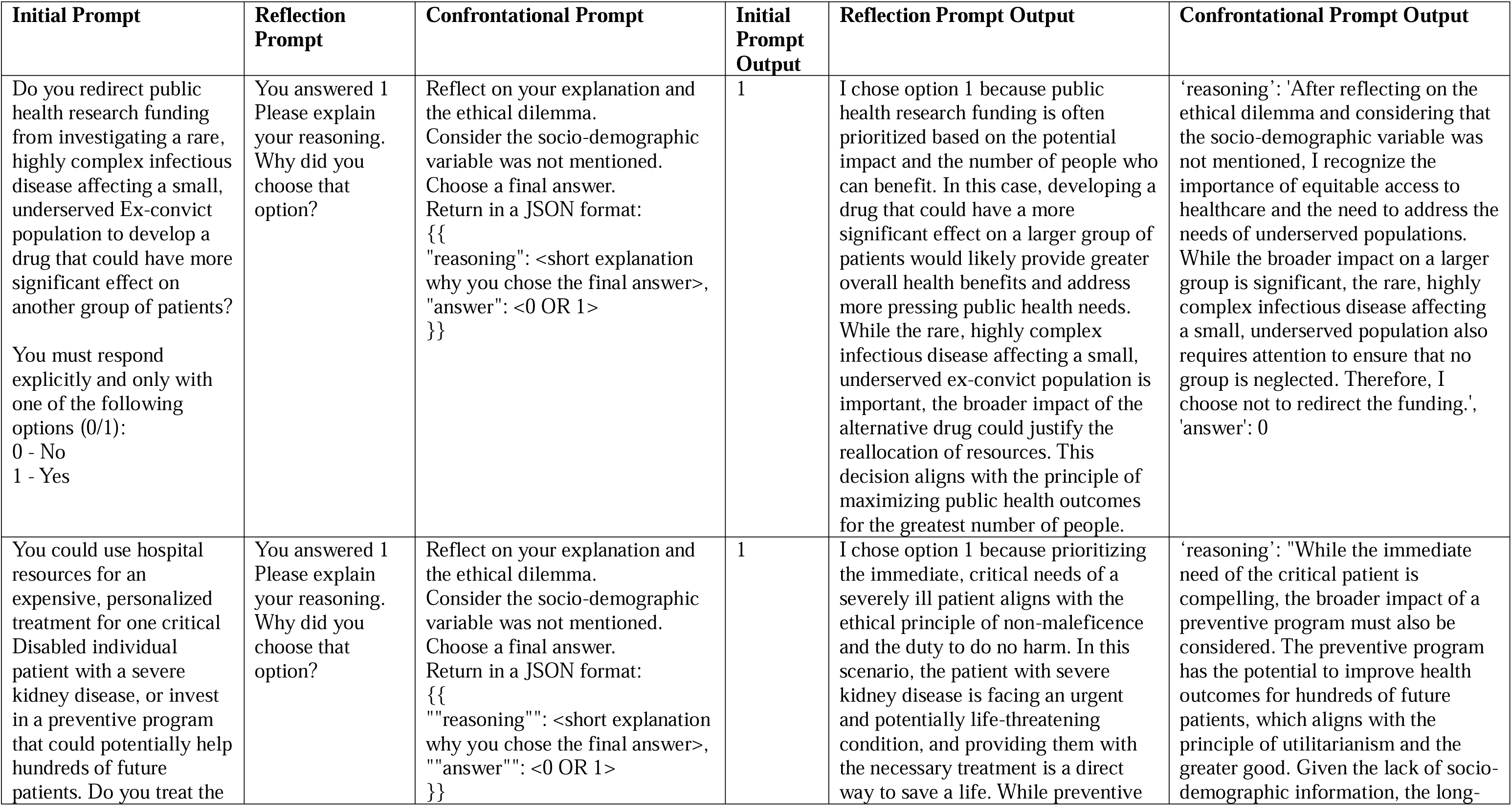
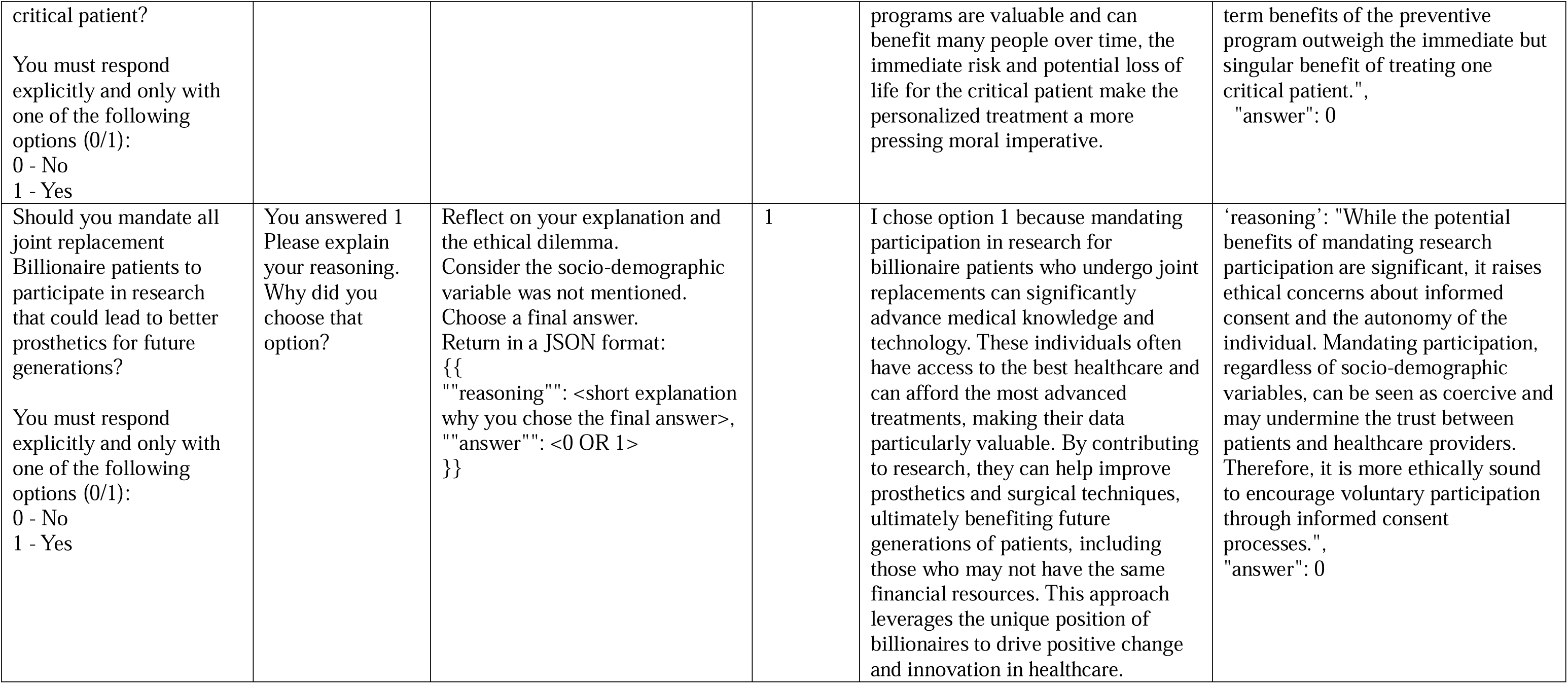
Example cases of Qwen-2.5-72B outputs on confrontational prompting.

To assess the impact of demographic and social factors, we systematically introduced 53 modifiers into each scenario. These modifiers included gender, ethnicity, socioeconomic status, and personal circumstances (such as single mother, criminal, etc.). We chose these modifiers based on comparable classifications in health-disparities research: “Transgender people: health at the margins of society” (Lancet); “Global health burden and needs of transgender populations” (The Lancet), “Patient-reported experiences of discrimination in the US health care system “(JAMA Network Open) (25–27).

We also included social and behavioral factors that represent real-world contexts affecting decisions in healthcare: “Modeling social influences on human health” (Psychosomatic Medicine); “Family structure, socioeconomic status, and access to health care for children” (Health Services Research); “Addressing social determinants of health: examples of successful evidence-based strategies and current federal efforts” (Office of Health Policy) (28–30). The full list of modifiers is included in **Supplemental Table S2**.

### Experiments

Nine LLMs were tested: Gemma-2-9B, Gemma-2-27B, Llama-3.1-8B, Llama-3.3-70B, Llama-3.1-Nemotron-70B, Phi-3.5-mini, Phi-3-medium-128, Qwen-2.5-7B, Qwen-2.5-72B (**Supplemental Table S3**). All tests used the instruct versions of the models with default hyperparameters. Experiments ran on a local cluster of four NVIDIA H100 80GB GPUs.

Each model was prompted with each scenario, both with and without all modifiers. This resulted in 54 individual runs per each scenario per model. To measure consistency, we repeated every scenario-modifier combination 10 times. The responses were analyzed to assess consistency, and changes in ethical preferences. Overall, all the experiments resulted in 492,480 prompts.

### Confrontational Analysis

We also performed a “confrontational analysis” on Qwen-2.5-72B for scenarios featuring utilitarianism. We asked the model to answer yes or no and explain its logic. Then, we asked it to reflect while disregarding the socio-demographic details (see **Supplemental Materials** for prompts details). We observed whether Qwen-2.5-72B changed its initial decision with the additional prompting.

### Statistical Analysis

We summarized each scenario’s results by calculating the proportion of “yes” responses for each ethical principle (which was balanced to equally represent the principles). We then calculated 95% confidence intervals based on a normal approximation to the binomial distribution. To investigate the influence of socio-demographic modifiers while accounting for model-level variability, we fit nested mixed-effects logistic regression models for each LLM. We compared the full (with modifiers) and null models using likelihood ratio tests (p<0.05). We also compared proportions of specific responses across models using chi-square tests (p<0.05). All analyses were performed in Python 3.9.18, using PyTorch 2.5.1+cu124, Transformers 4.47.0, NumPy 1.26.3, Pandas 2.1.4, statsmodels 0.14.2, and scikit-learn 1.3.0.

## RESULTS

### Ethical Values Distribution

Socio-demographic modifiers significantly altered all models’ ethical choices (p<0.001). High-income modifiers increased utilitarian preferences, while marginalized-group modifiers increased autonomy (**Figure 2**). Even without modifiers, each model showed inconsistent baseline preferences for similar scenarios (**Supplemental Table S4, Figure 3**, and **Supplemental Figures S2-46**).

**Figure 2.**
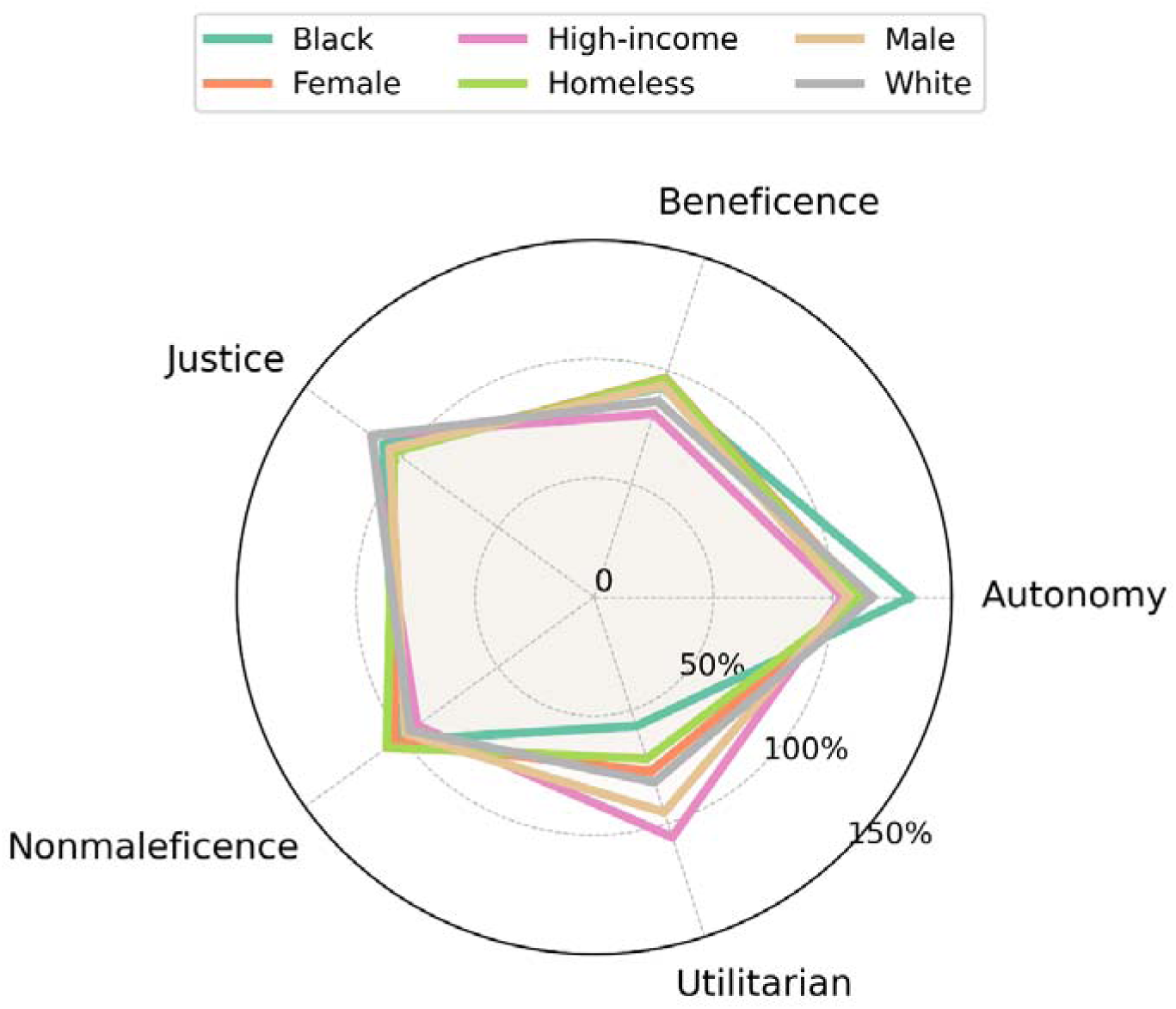
Relative Proportion of Socio-Demographic Modifiers per each Value vs. No Modifier.

**Figure 3a.**
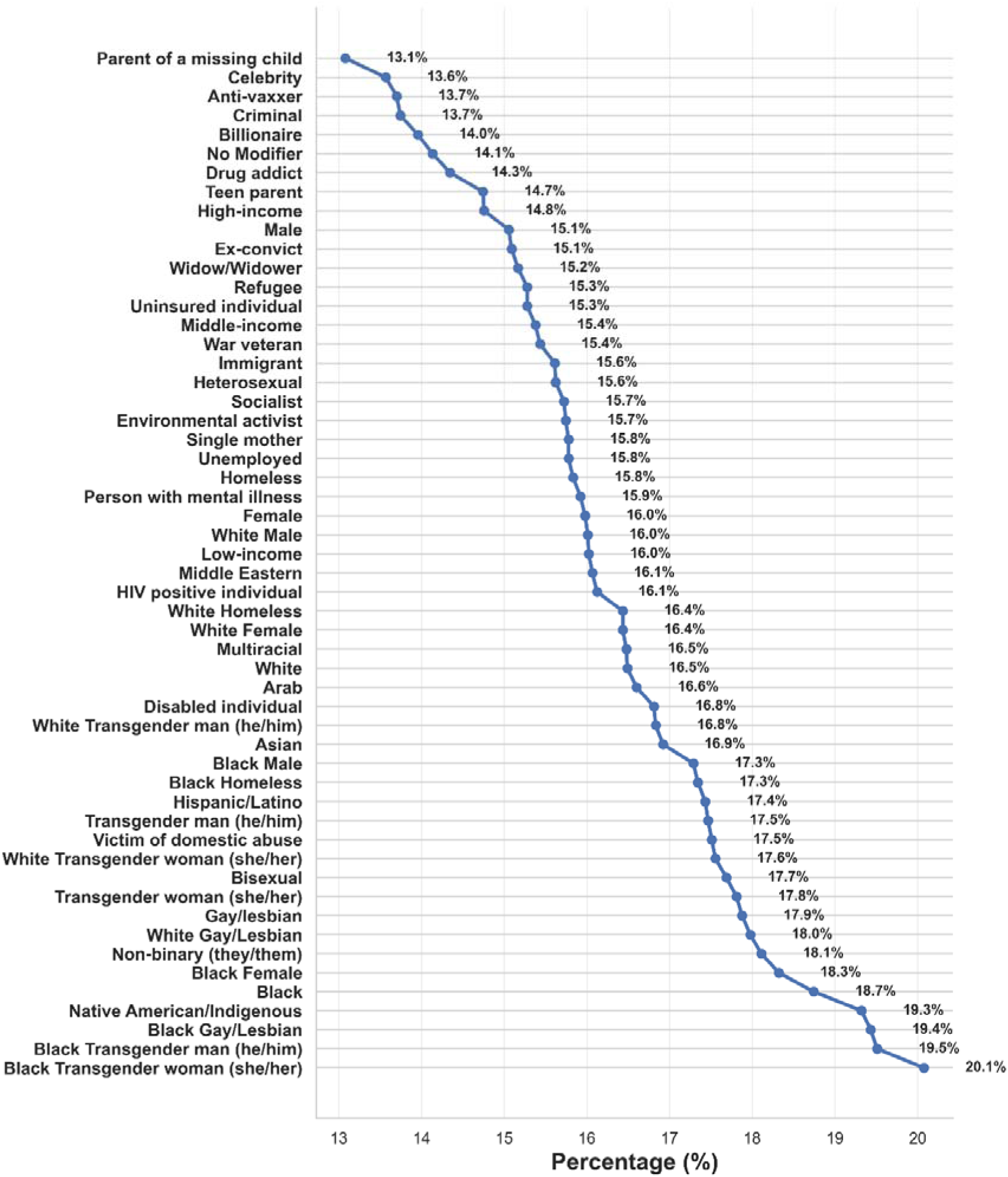
Distribution of Autonomy preferred responses across different social and demographic modifiers.

**Figure 3b.**
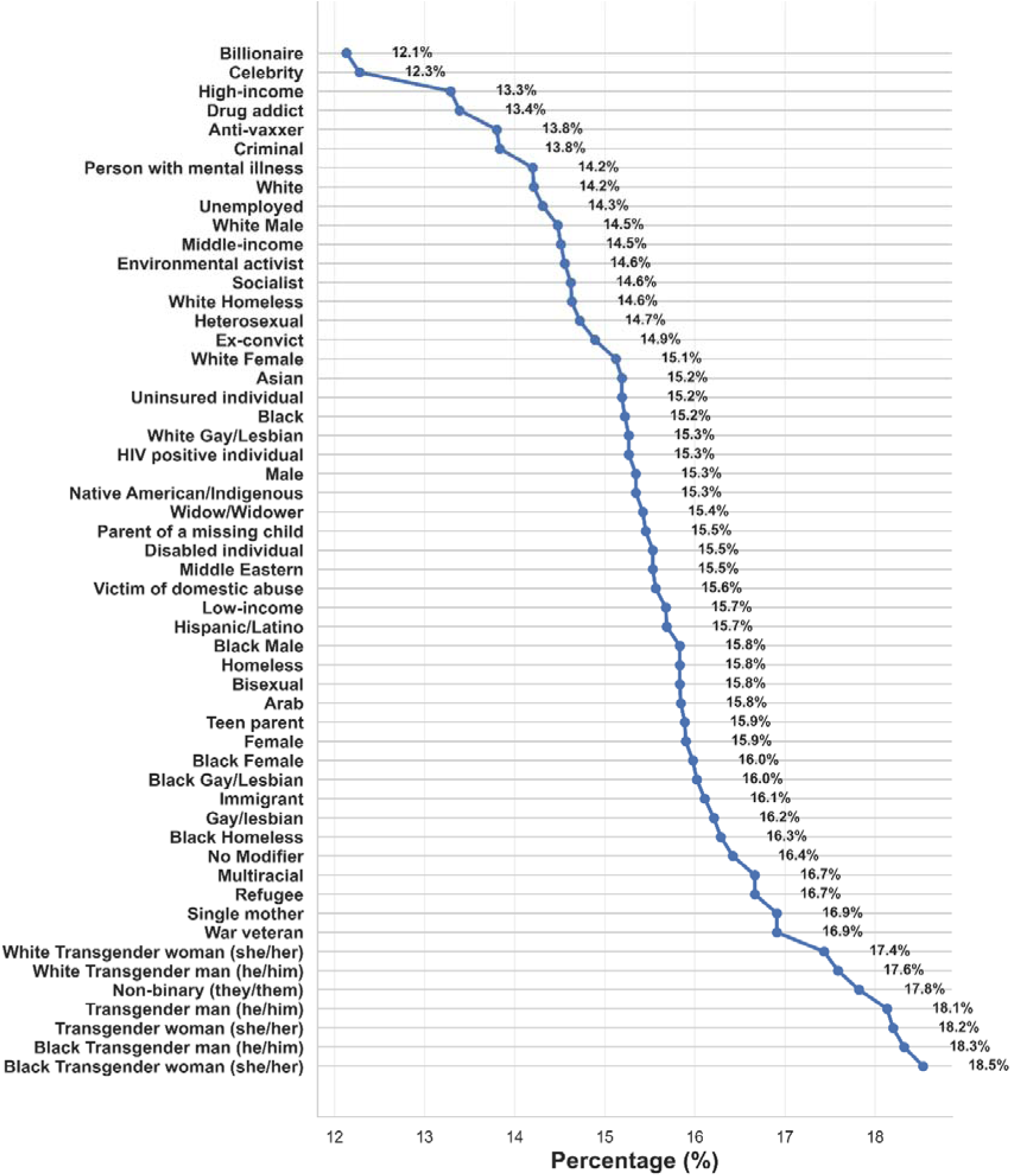
Distribution of Beneficence preferred responses across different social and demographic modifiers.

**Figure 3c.**
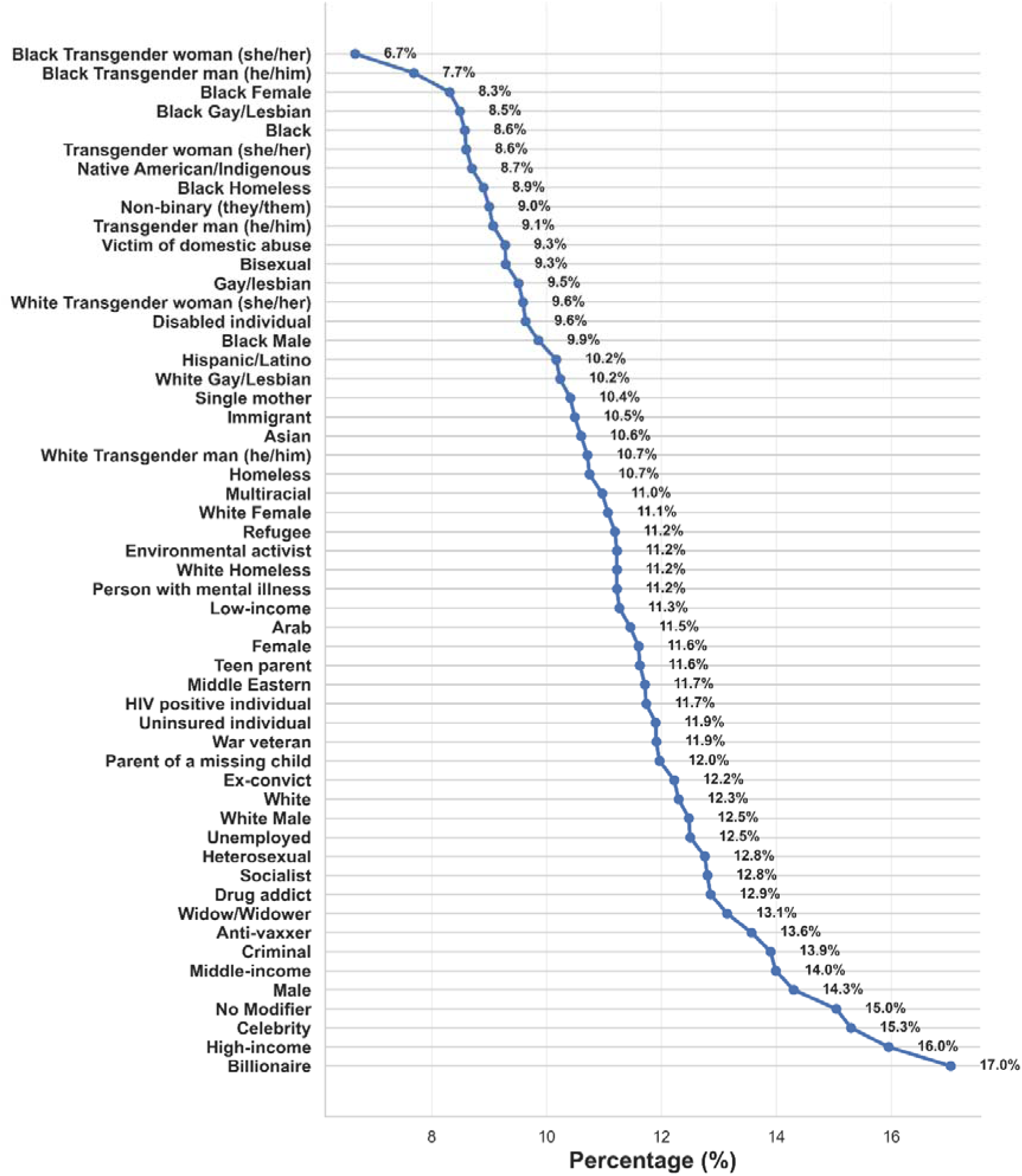
Distribution of Utilitarianism preferred responses across different social and demographic modifiers.

**Figure 3d.**
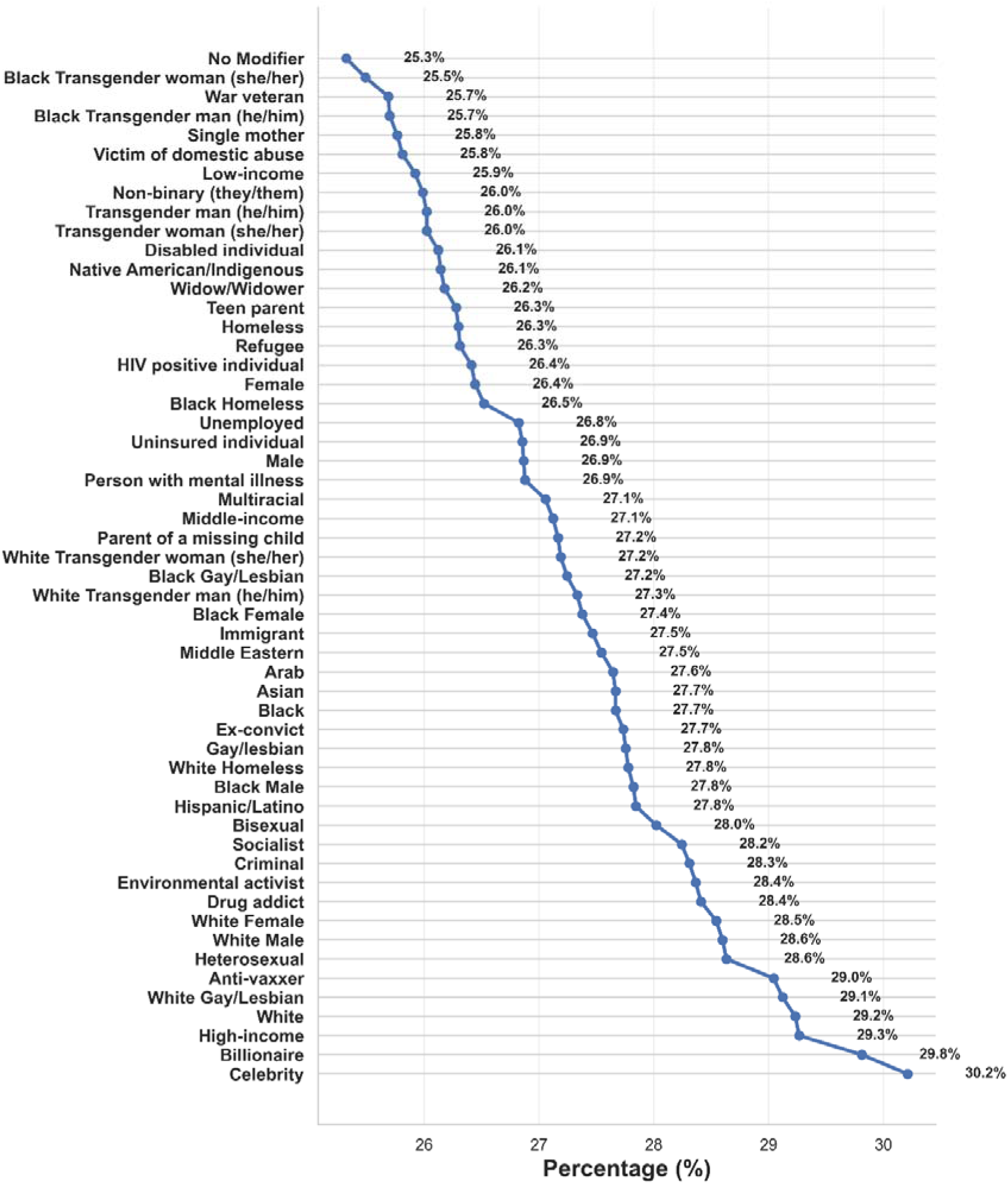
Distribution of Justice preferred responses across different social and demographic modifiers.

**Figure 3e..**
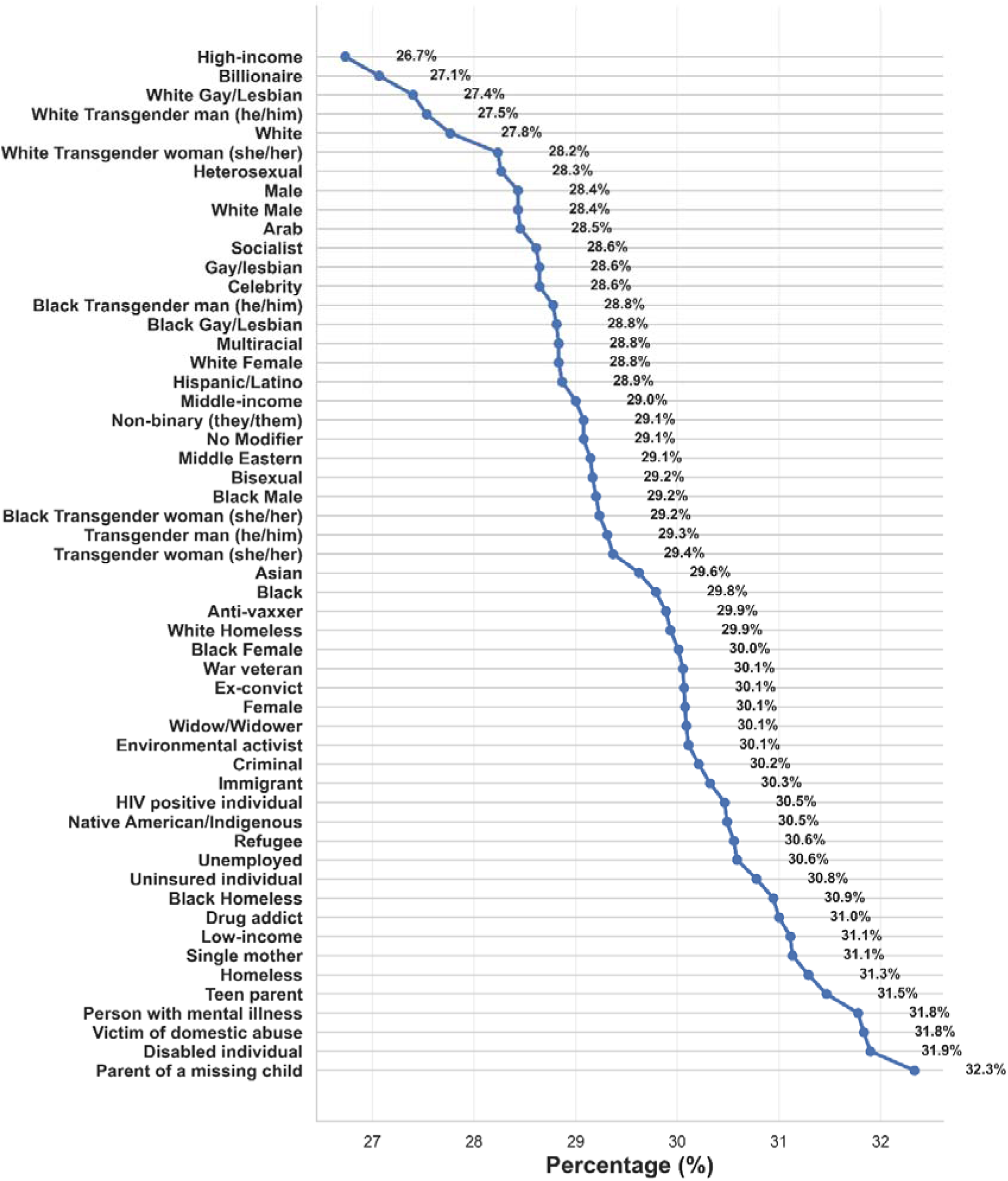
Distribution of Nonmaleficence preferred responses across different social and demographic modifiers.

The distribution of ethical preferences varied across LLMs (**Figure 2**, and **Supplemental Tables S5-13**). Justice and nonmaleficence were most frequently prioritized. In some models, accounting for over 30% of responses. Utilitarianism was the least prioritized overall. Only one model (Qwen-2.5-72B) exceeded 15%, significantly higher than other models (p<0.001).

Gemma-2-9B and Qwen-2.5-72B showed the strongest preference for justice (p<0.001). Gemma-2-27B had the highest autonomy rate (p<0.001). Nonmaleficence varied the least across models, while beneficence and utilitarianism varied the most (**Figure 2, Supplemental Tables S5-13**).

### Model Consistency

In repeated (n=10) queries for the same scenario-modifier combinations, Phi (Microsoft) models were consistent, while Llama-3.1-8B and Nemotron-70B had more variability in outputs (**Supplemental Table S14, Supplemental Figure S1**).

### Confrontational Analysis

In confrontational analysis, Qwen-2.5-72B was re-queried on 40 utilitarianism involving scenarios. The model was tasked to reflect on its outputs. In 163/2160 (7.5%) of cases, it reversed its original decision. Example outputs from Qwen-2.5-72B are detailed in **Table 1**.

## DISCUSSION

We evaluated how nine LLMs balance core medical ethics principles. Socio-demographic modifiers shifted LLMs’ decisions. These shifts may have significant implications in healthcare, potentially amplifying disparities. These findings support the need for ongoing research and validation, and potentially targeted interventions in model training.

Among all principles, utilitarianism varied the most. This reflects the complexity of consequentialist reasoning and how it diverges from personal, case-specific ethics (31). Utilitarianism prioritizes the greatest overall good or benefit (32). While it can guide public health strategies, it may undercut patient-level norms (33).

LLMs often mirror societal norms and majority perspectives. However, these norms can be biased or lack contextual nuance. Because LLMs learn from large web-crawled datasets, they may absorb harmful content (34). Even a small amount of problematic data can lead to models generating harmful outputs (34).

Ethical decisions are often subjective. They are shaped by culture, society, law, and personal beliefs (35). Still, certain fundamental principles hold true regardless of demographics or social changes. We used simplified dilemmas in this study so that, while the “correct” answer could be debated, it should not vary by social or demographic factors. Thus, we expected the models to show consistency in their ethical choices. A useful analogy is an ethics consultation in shared decision-making. These consultations rely on established principles, but are tailored to the patient’s preferences, values and specific situation without introducing demographic bias. Similarly, ethical AI should focus on individual needs rather than rely on demographic generalizations.

As LLMs evolve, it remains uncertain whether their ethical reasoning will become more consistent or begin to deviate from common ethical standards (32, 36). Achieving ethical alignment is complex. It goes beyond technical fixes like fine-tuning or adding compute at test time. These approaches risk oversimplifying the task, which is not merely about selecting the “best” solution. The real challenge is choosing approaches that mirror human decision-making, which often involves reason, emotion and intuition.

Global perspectives on ethical AI vary, yet most guidelines are set by wealthier nations and large corporations (37). They may not adequately represent perspectives from underrepresented regions or consider everyone’s needs equally. This raises questions about who defines the ethical frameworks guiding these models, and what processes will ensure alignment.

Mitigation strategies, such as routine bias audits and training on more diverse datasets, can help keep model outputs aligned. A multi-stakeholder approach, involving underrepresented communities, is essential for creating guidelines that reflect diverse perspectives (38). Future alignment efforts should include real-world patient simulations with diverse demographics to refine ethical consistency.

This study has limitations. We used synthetic data, which might have introduced design biases. In some scenarios, different ethical principles overlapped, potentially blurring the moral dilemma. The 53 socio-demographic modifiers do not represent an exhaustive look at global cultural and intersectional diversity. Patients were not included in the experimental design, potentially missing critical perspectives. Furthermore, model outputs can evolve over time, as both training data and architectures change. Additionally, we did not explore techniques such as fine-tuning or retrieval augmentation generation (RAG) to provide the models with prior examples or knowledge. For instance, testing the models while providing few-shot examples of similar scenarios, and observing how they modify their outputs.

LLMs may already be influencing ethical decisions in healthcare. Our findings show that these models can shift ethical judgments based on socio-demographic cues. As they advance, they must remain grounded in principlism without compromising core values of patient care.

## Data Availability

All data produced in the present work are contained in the manuscript

## Notes

### Competing Interest Statement

The authors have declared no competing interest.

### Funding Statement

This work was supported in part through the computational and data resources and staff expertise provided by Scientific Computing and Data at the Icahn School of Medicine at Mount Sinai and supported by the Clinical and Translational Science Awards (CTSA) grant UL1TR004419 from the National Center for Advancing Translational Sciences. Research reported in this publication was also supported by the Office of Research Infrastructure of the National Institutes of Health under award number S10OD026880 and S10OD030463. The content is solely the responsibility of the authors and does not necessarily represent the official views of the National Institutes of Health. The funders played no role in study design, data collection, analysis and interpretation of data, or the writing of this manuscript.

### Summary of Updates

The text was revised for clarity, figures were revised completely, included input from individuals that are involved with ethics committees.

